# Hepatitis B virus Seroprevalence Among Heterosexual Couples Attending Antenatal Care at Gulu Regional Referral Hospital, Uganda

**DOI:** 10.64898/2026.08.04.26359659

**Authors:** Patrick Otim, Felix Bongomin, Emmanuel Ochola

## Abstract

**Background:** Hepatitis B virus (HBV) is a global health concern and the leading cause of chronic liver disease, including liver cirrhosis and hepatocellular carcinoma. The Uganda Ministry of health recommends screening of HBV for all pregnant mothers during antenatal care (ANC). However, limited literature exists regarding couple testing and outcomes, particularly in Northern Uganda.

**Objective:** We determined HBV seroprevalence and associated factors among pregnant women and their partner attending ANC clinic of Gulu Regional Referral hospital (GRRH), Gulu, Uganda.

**Methods:** Between 1^st^ December 2023 and 28^th^ February 2024, a hospital-based, cross-sectional study was conducted among eligible pregnant women and their male partners attending ANC of GRRH, Gulu, Uganda. Every 12^th^ couple was enrolled following systematic random sampling; pretested questionnaire was administered by an interviewer. Blood samples were collected for HBV surface antigen (HBsAg) rapid test, which was performed according to manufacturer’s instructions. Quantitative data was analysed using logistic regression in STATA version 15 SE. P<0.05 was considered statistically significant.

**Results:** A total of 242 couple pairs were enrolled. Overall, 8.3% (n=40/484) participants were seropositive for HBsAg; A mong the males, Male partners (10.7%, n=26/242) and among pregnant women (5.8%, n=14/242). Sero-concordance 87.6% (212/242), (Male partners negative [-ve] and pregnant women [-ve]; 83.5%, n=202/242), (Male partners positive [+ve] and pregnant women [+ve]; 4.1%, n=10/242) and sero-discordance was 12.4% (30/242), (Male partners +ve and pregnant women -ve; 8.7%, n=21/242), (Male partners -ve and pregnant women +ve; 3.7%, n=9/242). Factors associated with positive HBsAg were, being male (adjusted odds ratio [aOR]: 2.45, 95% Confidence Interval [CI], 1.15 - 5.21, p=0.02), daily alcohol consumption (aOR: 4.73, 95% CI: 1.40 -16.04, p=0.01), and no prior vaccination against HBV (aOR: 12.01, 95% CI: 4.07 - 35.65, p<0.001).

**Conclusion:** There is a high HBsAg seropositivity rate, particularly among male partners of pregnant women. There is an urgent need to have couples who come for ANC to be screened for HBV and to strengthen and sensitise young people for premarital HBV screening.

## INTRODUCTION

Hepatitis B virus (HBV) is a viral infection of major global public health importance, capable of causing both acute and chronic liver disease and leading to severe complications such as hepatocellular carcinoma (HCC), cirrhosis, and fulminant hepatic failure. Acute HBV infection commonly presents with symptoms including jaundice, dark urine, abdominal pain, anorexia, fatigue, nausea, and vomiting. In contrast, chronic HBV infection may remain asymptomatic for many years but can result in progressive liver damage over time, making it a major health priority, particularly in developing countries [1–4].

The World Health Organization (WHO) estimates that approximately 254 million people were living with chronic hepatitis B globally by 2022 corresponding to about 3.2% of the world population. In the same year, viral hepatitis caused an estimated 1.3 million deaths, making it one of the few communicable diseases for which mortality is increasing. Of these deaths, hepatitis B accounted for about 1.1 million, while hepatitis C contributed approximately 240,000 deaths [1].

HBV is transmitted through multiple routes. In highly endemic settings, mother-to-child transmission during childbirth is a major mode of transmission, while horizontal transmission through contact with infected blood during early childhood also contributes substantially [5]. Evidence suggests that up to 40% of chronic HBV infections in sub-Saharan Africa are acquired during early childhood [6]. Additionally, in utero transmission during pregnancy has been reported and may be associated with adverse pregnancy outcomes such as preterm delivery [7]. Other transmission routes include infected male patterner, exposure to infected body fluids through sexual contact, unsafe injections, needle-stick injuries, tattooing, piercing, and sharing contaminated syringes in healthcare settings or among people who inject drugs [8].

Infants born to mothers who are positive for both hepatitis B surface antigen (HBsAg) and hepatitis B e antigen (HBeAg) have a 70–90% risk of acquiring perinatal HBV infection, whereas infants born to mothers who are HBsAg-positive, but HBeAg-negative have an approximately 25% risk. Among infants infected perinatally, 85–90% progress to chronic infection, facilitating ongoing horizontal transmission. Approximately 25% of individuals with chronic HBV infection eventually die from HCC or complications of liver cirrhosis [9].

Several studies across Africa report consistently high HBV prevalence among pregnant women. In Nigeria, a cross-sectional study among pregnant women and their male partners participating in a baby shower program reported an HBV prevalence of 10.9%, indicating a substantial disease burden [10]. In The Gambia, screening of 423 pregnant women attending antenatal care revealed an HBV seroprevalence of 9.2%, with significantly lower prevalence among vaccinated women 2.3% compared with unvaccinated women 13.7% [11].

In Uganda, more recent studies conducted at Kyazanga Health Centre IV and Mulago National Referral Hospital reported HBV prevalence rates of 2.8% and 2.9%, respectively, among pregnant women, falling within the WHO-defined intermediate endemicity range of 2–8% [12] [13]. However, an earlier cross-sectional study in 2014 conducted in two hospitals in Northern Uganda reported a much higher prevalence of 11.8% among pregnant women, suggesting a persistently high burden in this region where vertical and early horizontal transmission remain predominant [14].

Male partner involvement in antenatal care (ANC) significantly improves maternal and neonatal health outcomes. Beyond emotional and financial support, it enables earlier detection of pregnancy complications, increases uptake of key services (e.g., skilled delivery, postnatal care, and couple-based HBV/HIV testing), and reduces risks to both partners and the newborn such as mother-to-child transmission, undiagnosed maternal conditions, and adverse neonatal outcomes. For male partners, participation in couple testing supports early detection and management of their own infections, lowering risks of chronic disease and transmission. Limited male involvement, however, can delay danger sign recognition, reduce service uptake, and increase preventable morbidity and mortality for mother, father, and child.[15–17].

Despite the high burden of HBV in Uganda there is limited seroprevalence and infection patterns among pregnant women and their male partners attending ANC, particularly regarding concordance and discordance with in couples. Male partners remain significantly under screened in ANC settings, and there is a notable lack of region-specific data from Uganda and northern Uganda. The knowledge gap hinders the development of targeted couple-based intervention for HBV prevention and control in high prevalence setting.

Therefore, this study aimed to determine the seroprevalence of HBV among pregnant women and their male partners, assess concordance and discordance of HBV infection within couples, and identify associated factors among couples attending ANC at Gulu Regional Referral Hospital (GRRH) in northern Uganda.

## METHODS

### Study Design and setting

This was a facility-based cross-sectional study employing a quantitative approach between 1^st^ December 2023 and 28^th^ February 2024. The study was conducted at the ANC of GRRH, located in Gulu City, Northern Uganda. GRRH is a tertiary referral facility serving surrounding and neighbouring districts and has an approximate capacity of 250 beds. The antenatal clinic operates Monday to Friday from 8:00 am to 5:00 pm (excluding public holidays) and receives an average of 50–70 antenatal visits per day.

### Study Population

The study population consisted of pregnant women and their male partners attending ANC services in Northern Uganda, with the accessible population being those who attended the GRRH ANC. The primary study unit was defined as a couple (a pregnant woman and her male partner) attending ANC together. Eligibility required that both members of the pair provide written informed consent, with no age restrictions applied. Pregnant women presenting with medical or obstetric emergencies requiring immediate care were excluded from participation.

### Study Variables

The dependent variable for this study was the HBsAg serostatus, assessed among both pregnant women and their male partners. Independent variables included sociodemographic characteristics such as age, sex, marital status, educational level. Obstetric history factors comprised gravidity, place of previous delivery, and history of abortion. Sexual history variables included HIV status, history of sexually transmitted infections, and having multiple sexual partners. Lifestyle factors such as body piercing, tattoos, alcohol consumption, and substance use were also assessed, along with medical history elements including blood transfusion, road traffic accident involvement, and HBV vaccination status.

### Sample Size Estimation

Sample size was calculated using the Kish–Leslie (1965) formula for estimating a single population proportion. 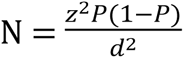

N is the required sample size. Z is the Z score which was set at 1.96 for 95%. d. is the margin of error at 5% and the P is the prevalence of a known population.

Three different prevalence estimates from previous studies were used to ensure adequate power for all study objectives

Using a prevalence of 10.9% among pregnant women and partners (Talla et al., 2021) the minimum sample size was 150 participants.

A couple-level prevalence of 19.5%, produced sample size was 242 couples

prevalence 11.7% for associated factors gave a sample size was 159 couples.

The largest sample size 242 couples (corresponding to 484 individual) were adopted to provide sufficient statistical power across all objectives. No adjustment was made for non-response as the study achieved full recruitment of the target sample with no refusals.

### Sampling Procedure

Systematic random sampling was used to recruit 242 couples. The sampling frame was the daily ANC registration logbook with approximately 50–70 couples attending daily on working days except public holidays and weekends, yielding about 1,200 couple visits per month and a sampling interval of K=12 was applied. To get the kth couple, we started counting from the first couple registered in the ANC registration book and the 12^th^ couple registered was recruited and every 12^th^ couple. This approach was maintained over 60 working days, with approximately 5 couples enrolled per day to achieve the final sample, Eligibility screening was conducted by trained research assistants.

### Data Collection Procedures

Data was collected using a pretested structured questionnaire administered by trained research assistants. Participants then jointly received pre-test counselling, provided finger-prick blood samples and received post-test counselling after results disclosure. Questionnaires were checked daily for completeness and securely stored. The procedure of counselling followed a structured, couple friendly approach in a private, non-judgmental environment (room). The pretest included rapport building, assessment of knowledge and risk factors, explanation of possible results (concordant negative, concordant positive, or discordant), and the testing process. The participants were offered the choice of receiving results separately or together. In discordant cases, results were first disclosed separately to each partner. Partners only learned each other’s Sero status after they consented to mutual disclosure. Positive partners were counselled and referred for further evaluation and care (in the chronic care clinic). Negative partners received counselling on prevention strategies, including vaccination and safety planning.

### Laboratory Procedures

Capillary blood samples were collected by trained licensed laboratory technician using standard infection prevention and control procedures with single use lancets. All samples were of adequate quality with no visible haemolysis, resulting in no exclusions or invalid results. HBsAg testing was performed immediately on site using the SD BIOLINE HBsAg WB rapid test kit (Abbott Korea Ltd., Seoul, South Korea). The manufacture reports 100% sensitivity and specificity compared to ELISA consistent with other studies [18]. In this field study with finger prick samples, internal quality controls were valid for all tests and results were read after 20 minutes according to manufacturer instructions. Participants received their results from the counsellor where post-test counselling was done.

### Data Management and Analysis

Data were entered into Microsoft Excel and exported to STATA version 15 SE for analysis. Descriptive statistics were used to summarize participant characteristics and HBV seroprevalence. Prevalence was calculated as:

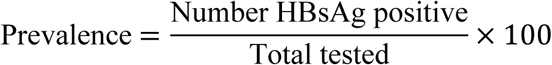

Couple-level concordance and discordance were computed as:

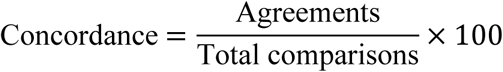

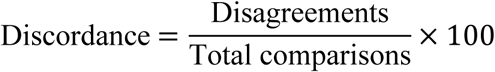

Bivariate analysis using chi-square tests and crude odds ratios identified variables associated with HBV seropositivity. Variables with p < 0.05 were entered into multivariable logistic regression to determine independent predictors of HBV infection and couple concordance/discordance. Model selection used forward stepwise regression with assessment of multicollinearity and goodness-of-fit.

### Ethical Considerations

Ethical approval was obtained from Gulu University Research Ethics Committee (GUREC-2023-598) and administrative clearance from GRRH. Written informed consent was obtained from all participants. Confidentiality and privacy were maintained using unique identifiers. Participation was voluntary, and participants could withdraw at any time. Individuals who tested positive were counselled and referred for appropriate care.

## RESULTS

### Characteristics of the Study Participants

During the study period, 484 participants were randomly selected, comprising 242 couples (Male partner and pregnant woman). The mean age of all participants was 28.91 years (standard deviation= 7.58). Most had secondary education 50.8%(n=246), were in monogamous marriages 76.7%(n=371), and were from the Acholi tribe 84.9%(n=411). In terms of occupation, elementary roles 26.0%(n=126) and services/sales 20.2% were most common. Catholics formed the largest religious group 46.3%(n=224). A majority earned under 1 million UGX monthly 91.8% (n=223) and 86.2%(n=417) were aware of HBV. Among recent mothers, 95.3%(n=142) delivered in health facilities. Most women 52.1%(n=126) had 1-3 children, and 24.8%(n=60) reported a history of abortion. Few had recent sexually transmitted infections (STIs) 7.4%(n=36) or road traffic accidents 11.4%(n=55), and 98.8%(n=478) had never received a blood transfusion. The majority were HIV-negative 94.6% (n=458), 51.2%(n=248) were vaccinated against HBV, and 11.6%(n=56) had piercings or tattoos, **Table 1**.

**Table 1.** Characteristics of the Study Participants.

| **Variable** | **Frequency** | **Percentage (%)** |
| --- | --- | --- |
| **Age (mean)** | 28.91 (17-60) |  |
| **Sex** |  |  |
| Female | 242 | 50.0 |
| Male | 242 | 50.0 |
| **Education levels** |  |  |
| Primary | 159 | 32.9 |
| Secondary | 246 | 50.8 |
| Tertiary | 79 | 16.3 |
| **Type of Marriage** |  |  |
| Monogamous | 371 | 76.7 |
| Polygamous | 113 | 23.3 |
| **Tribe** |  |  |
| Acholi | 411 | 84.9 |
| Lango | 21 | 4.4 |
| Others | 52 | 10.7 |
| **Occupation** |  |  |
| Professional | 63 | 13.0 |
| Services and sales | 98 | 20.2 |
| Skilled agricultural, fish | 95 | 19.6 |
| Craft and related | 56 | 11.6 |
| Elementary occupation | 126 | 26.0 |
| Armed forces | 23 | 4.8 |
| Others | 23 | 4.8 |
| **Religion** |  |  |
| Anglican | 154 | 31.8 |
| Catholic | 224 | 46.3 |
| Born again | 20 | 4.1 |
| Muslim | 79 | 16.3 |
| Others | 7 | 1.5 |
| **Monthly income** |  |  |
| Less than 1 million UGX | 223 | 91.8 |
| Above 1 million UGX | 20 | 8.2 |
| **Aware about HBV** |  |  |
| No | 67 | 13.8 |
| Yes | 417 | 86.2 |
| **\* Place of last birth** |  |  |
| (n = 149) |  |  |
| Health facility | 142 | 95.3 |
| Traditional birth attendance | 7 | 4.7 |
| **Parity (n = 242)** |  |  |
| 0 | 91 | 37.6 |
| 1-3 children | 126 | 52.1 |
| ≥ 4 children | 25 | 10.3 |
| **History of abortion** |  |  |
| (n = 242) |  |  |
| No | 182 | 75.2 |
| Yes | 60 | 24.8 |
| **STI in past three months** |  |  |
| No | 448 | 92.6 |
| Yes | 36 | 7.4 |
| **Alcohol use** |  |  |
| No | 384 | 79.3 |
| Yes | 100 | 20.7 |
| **Frequency of alcohol intake** |  |  |
| Do not take | 386 | 79.7 |
| Occasional | 72 | 14.9 |
| Daily | 26 | 5.4 |
| **History of blood transfusion** |  |  |
| Never transfused | 478 | 98.8 |
| Ever transfused | 6 | 1.2 |
| **History of RTA** |  |  |
| No | 429 | 88.6 |
| Yes | 55 | 11.4 |
| **HIV status** |  |  |
| Negative | 458 | 94.6 |
| Positive | 26 | 5.4 |
| **Vaccination against HBV** |  |  |
| No | 236 | 48.8 |
| Yes | 248 | 51.2 |
| **Having piercings/tattoo** |  |  |
| No | 428 | 88.4 |
| Yes | 56 | 11.6 |

*\*The place of last birth includes the primipara and Multipara mothers only (n=149)*

### Hepatitis B Sero Concordance and Discordance

HBsAg discordance rate was 12.4% (30/242), 8.7% (n=21/242) among couples where the male partner is HBsAg positive and the pregnant woman is negative while a discordance rate of 3.7% (n=9/242) was observed among couples where the male partner was HBsAg negative and the female was HBsAg positive. The concordance rate was 87.6% (n=212/242), 4.1% (n=10/242) among couples where both partners were HBsAg positive, and 83.5% (n = 202/242) among couples where both partners were HBsAg negative, **Figure 1**.

**Figure 1.**
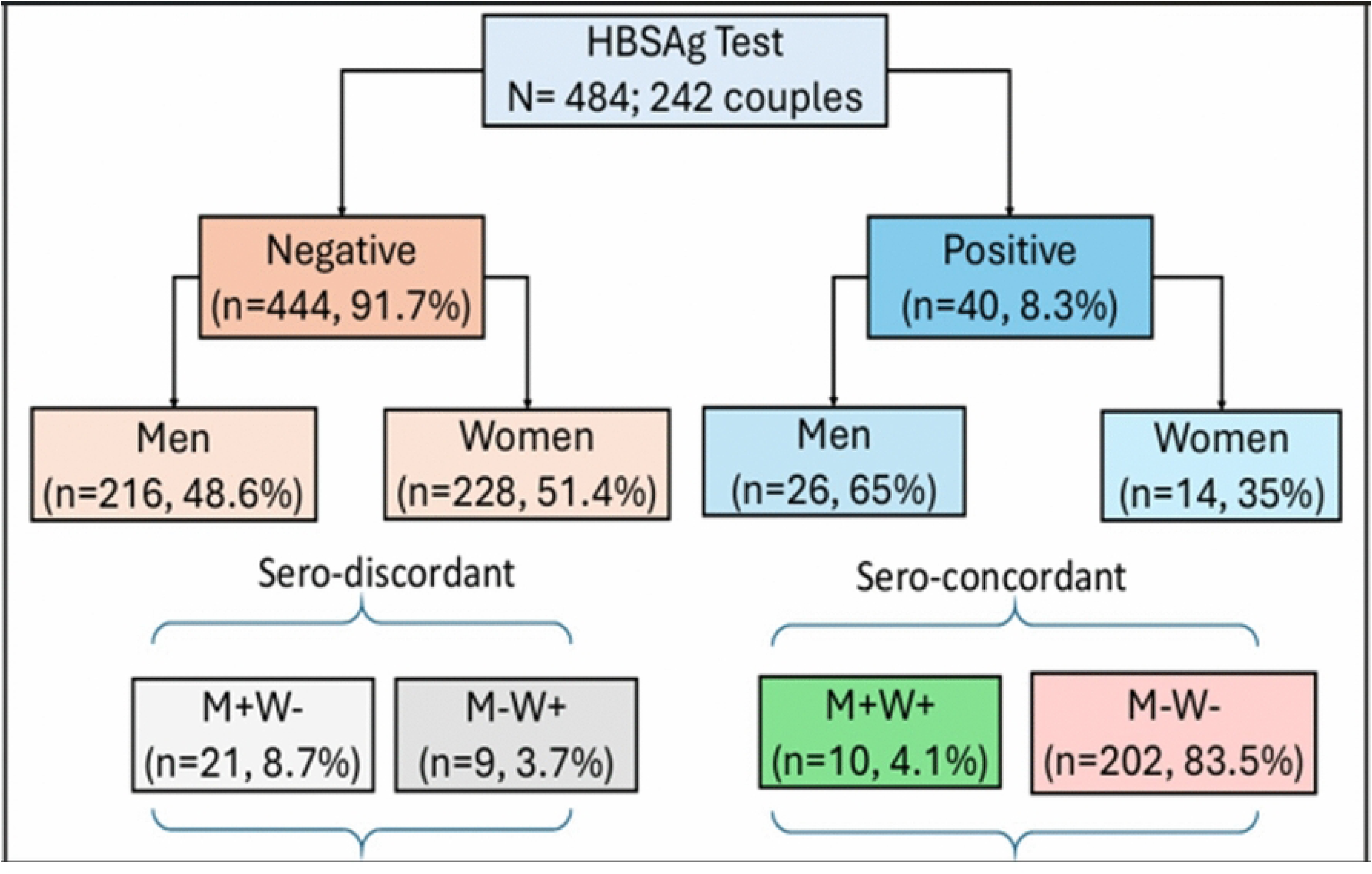
Hepatitis B surface antigen Sero-Concordance and Sero-Discordance.

### Bivariate analysis of factors associated with hepatitis B seropositivity

Factors statistically significantly associated with a positive HBsAg tests were ; being male (OR=1.96 95% CI: 1.00–3.85, p=0.05), Prior vaccination against HBV (OR=10.98, 95% CI: 3.84 – 31.37, p=0.001), daily alcohol consumption (OR:3.44 95% CI: 1.29–9.18, p=0.01), and receiving blood transfusion (OR=5.79, 95% CI: 1.03–32.64, p=0.05). **Table 2**

**Table 2.**
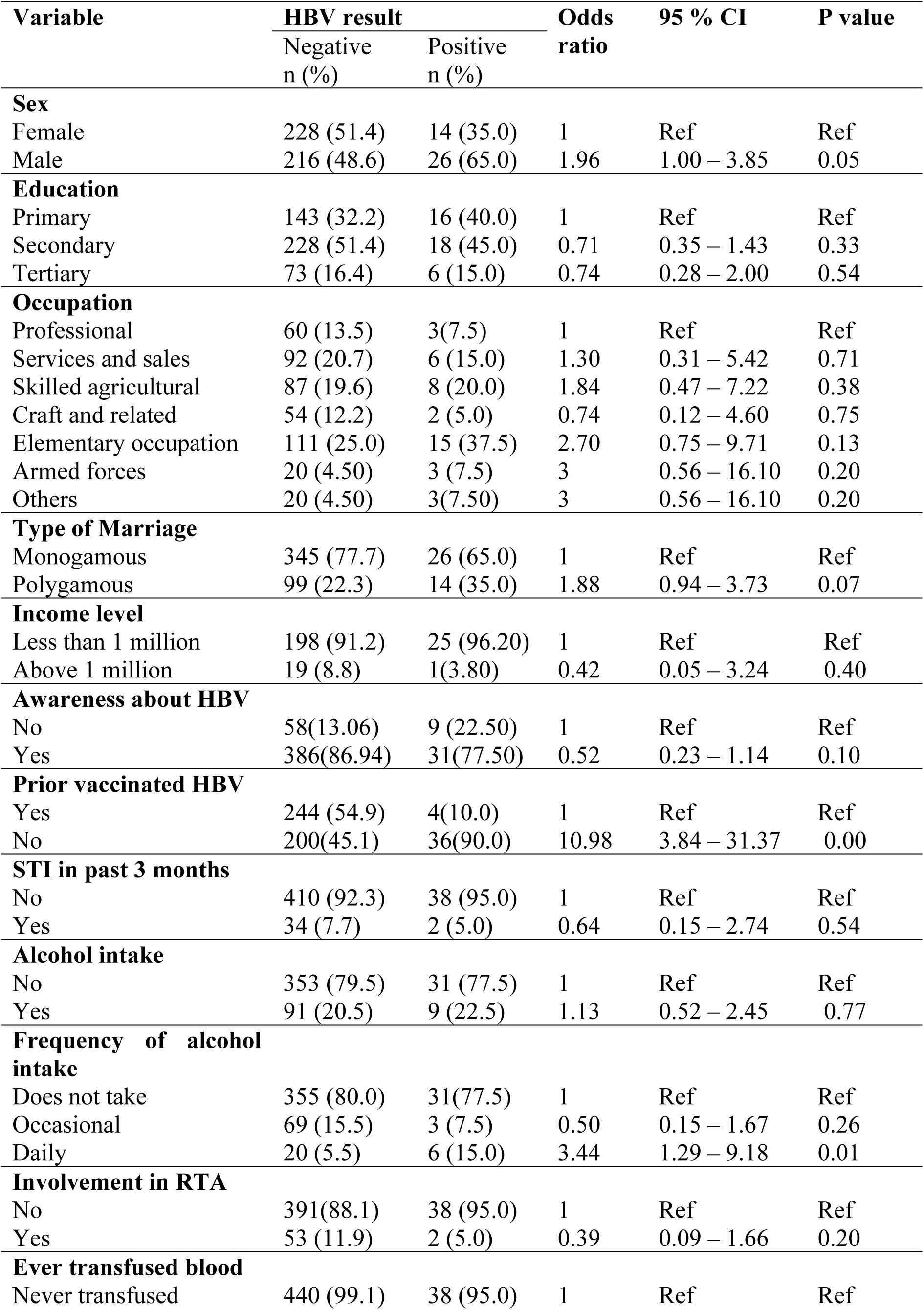
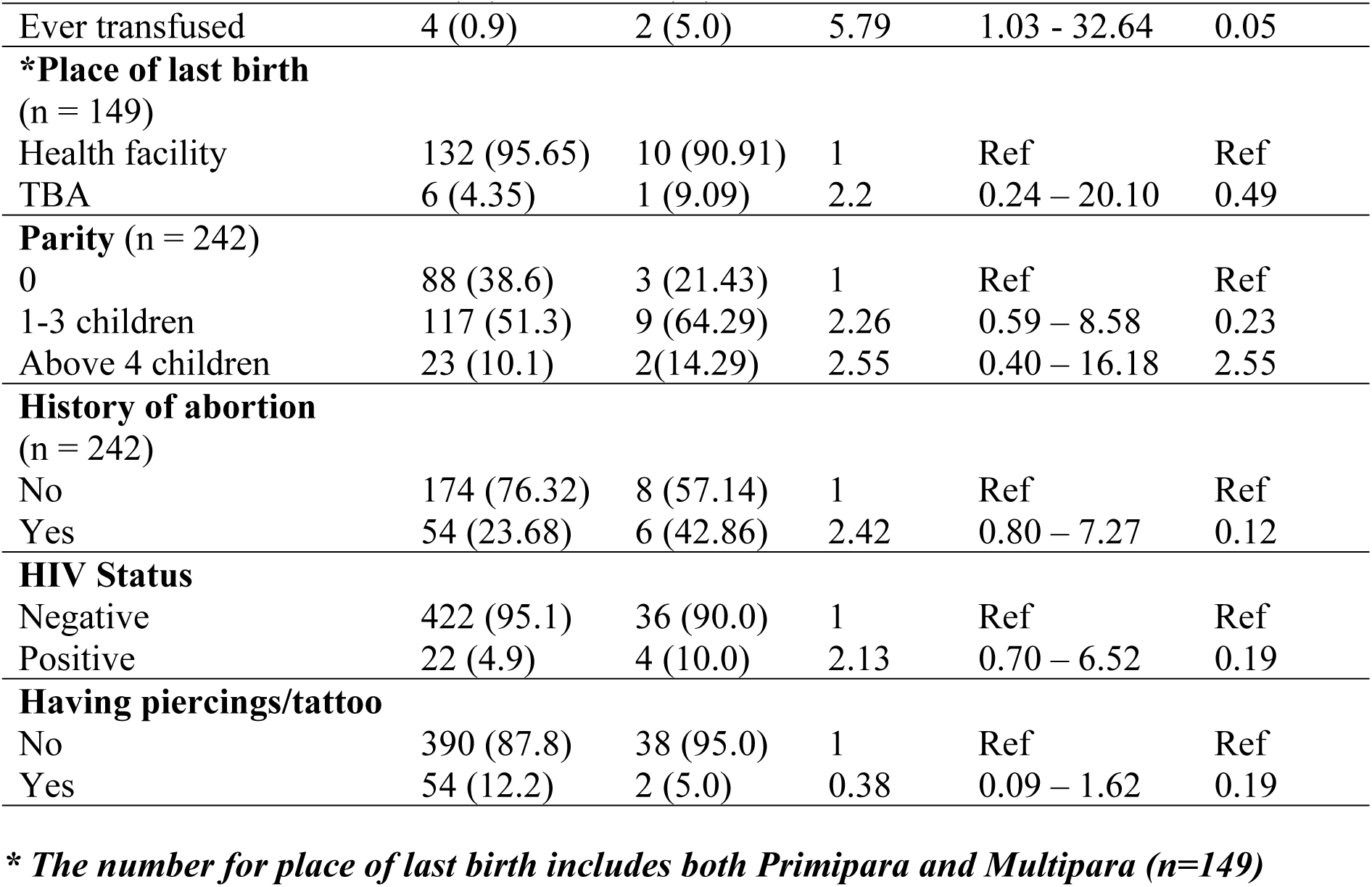
Bivariate analysis of factors associated with hepatitis B seropositivity.

### Multivariable analysis of factors independently associated with Hepatitis B positivity

The factors that were independently associated with Hepatitis B positivity were; Male gender (aOR =2.45, 95% CI: 1.15 – 5.21 p=0.02), No prior vaccination against HBV (aOR: 12.05, 95% CI: 4.07 - 35.65, p<0.001), Daily Alcohol consumption (aOR=4.73, 95% CI: 1.40 –16.04 p=0.01) and being involved in road traffic accident(aOR=0.2, 95% CI: 0.04 – 1.02 p=0.05) **Table 3**.

**Table 3.**
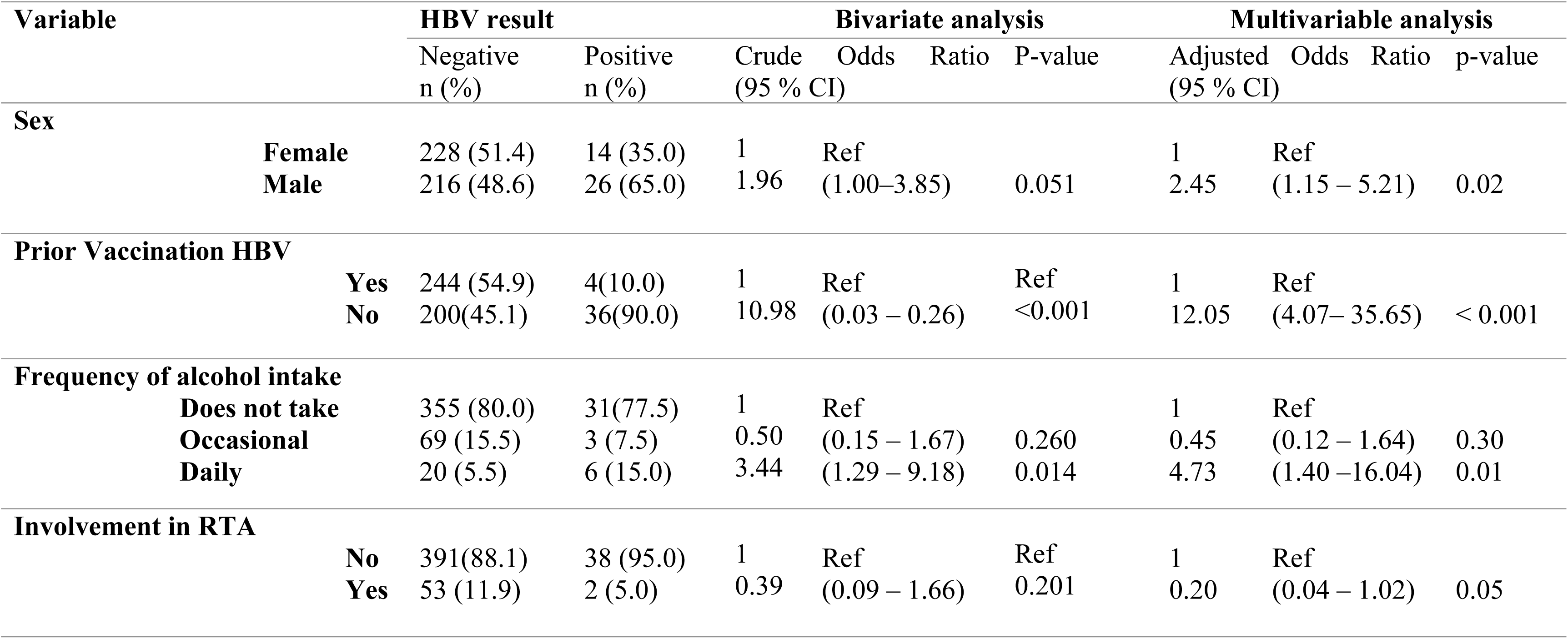
Multivariable analysis of factors independently associated with Hepatitis B positivity.

## DISCUSSION

In this study, we found an overall HBV seroprevalence of 8.3% among pregnant women and their male partners attending ANC at GRRH. HBV positivity was significantly higher among male partners than among pregnant women, indicating a male predominance of infection within couples. This pattern suggests that male partners may play a substantial role in sustaining HBV transmission within households and contributing to both horizontal and perinatal transmission.

Our findings are consistent with previous studies from Uganda reporting comparable HBsAg seroprevalence and similar sex differentials. A national survey[19]reported an overall HBV seroprevalence of 10.3%, with a higher prevalence among males 11.8% than females 9.1%, underscoring persistent male predominance at the national level. In contrast, a community-based study [20] reported a higher prevalence of 17.2%(n=123/717), which may reflect differences in population characteristics, and the timing of data collection relative to the scale-up of HBV vaccination in contrast our study was done on pregnant women and their male partners.

However, our prevalence of 5.8% among the pregnant women in notably lower that the 11.8% reported by Bayo et al in the same region among ANC attendees. This difference cannot be straightforwardly attributed to the scale up of infant HBV vaccination, which was introduced into Uganda’s expanded program on immunization in 2002. Given the mean age of the study participants of 28.9 years, majority of the participants were not eligible to routine childhood immunization however government introduced targeted adult vaccination programs starting with health workers in 2011 and expanding to the general population in 2016 following testing. Direct protective effects from routine infant vaccination would have been minimal in this cohort, however some participants could have received at least one dose during these later adult vaccination campaigns.

The observed difference may be attributed to several factors including, selection bias arising from recruiting only couples attending ANC together who may differ in socio-economic status and risk behaviours from the ANC population and possible temporal changes in transmission dynamics, blood safety or community awareness in post conflict Northern Uganda.

Other studies in Uganda provide additional context. Investigations among medical students and healthcare workers have reported prevalence estimates ranging from 9% to 11%, likely reflecting occupational exposure and increased risk in clinical environments. Nevertheless, these estimates remain broadly comparable to those observed in the general population, aligning with our findings [21,22]. In northern Uganda, Chiesa et al. reported an HBV prevalence of 7.9% among individuals co-infected with HIV, with a higher prevalence among males 11.7% than females 6.8% [23]. Despite the distinct characteristics of this subgroup, the similarity in prevalence to our study highlights the persistently high HBV burden in the region.

Globally, the WHO estimates that the African Region accounts for approximately 63% of new viral hepatitis infections, and the pooled prevalence of HBV among pregnant women in Africa is estimated at about 4.2%. Studies from West Africa report male predominance within couples, with higher prevalence among male partners than female partners. For example, Onakewhor *et al*., [24] reported HBV prevalence of 6.8% among male partners and 4.3% among female partners, while in Nigeria, Talla *et. al*. reported a seroprevalence of 10.9% among couples, with higher positivity among male partners 11.9% than pregnant females 10.2% [10]. Similar sex differentials have been documented in Asia, including China and Iraq [25–28]. Although absolute prevalence varies widely across settings, the consistent observation of higher HBV prevalence among males suggests that behavioural, occupational, and biological factors may contribute to sex-specific vulnerability, with important implications for household-level transmission.

We observed that most couples were concordantly HBsAg-negative 83.5%, while 4.1% were concordantly HBsAg-positive. However, a notable proportion of couples were sero-discordant 12.4%, with 8.7% comprising an HBsAg-positive male partner and HBsAg-negative pregnant woman, and 3.7% comprising an HBsAg-positive pregnant woman and HBsAg-negative male partner [10] These discordant partnerships represent a critical risk group for both horizontal transmission between partners and vertical transmission to infants.

The proportion of HBV cases among exposed couples was 12% compared with 8% among unexposed couples, yielding an absolute difference of 5%. The exact odds ratio of 2.33 indicates that individuals with an HBV infected partner had more than twice the odds of being HBV infected themselves compared to those without such exposure. These findings provide quantitative evidence of intra-couple transmission risk and support the importance of couple-based screening and prevention strategies.

Comparable patterns have been reported elsewhere. Studies in Nigeria documented that most couples were concordantly negative, with smaller proportions of concordant positive and discordant pairs [10,29]. In China, high concordant-negative rates and low concordant-positive rates have been attributed to long-standing universal childhood vaccination programs [27,28,30]. In Uganda, although HBV vaccination has been scaled up, gaps remain in adult vaccination coverage[31], which may explain the persistence of discordant and concordant positive couples. Together, these findings highlight the need for routine partner testing, counselling, and vaccination within antenatal care services.

Male sex was independently associated with HBV infection, with males having more than twice the odds of testing HBsAg-positive compared with females. Similar findings have been reported in Nigeria, Thailand, and China,[10,28,30,32–34]where male partners disproportionately contribute to discordant couple results. These observations underscore the importance of prioritizing male partner testing and linkage to care within ANC and partner-invitation programs.

No prior vaccination was strongly associated with HBV sero-positivity. Participants with no prior vaccination had over 12 times higher odds of testing positive for HBV.. This finding is consistent with the well-documented efficacy of hepatitis B vaccination in preventing infection and interrupting transmission [35]. Although Uganda introduced universal infant vaccination in 2002, adult catch-up coverage remains suboptimal. Our results strongly support immediate implementation of “test and vaccinate” strategies at ANC clinics, including on site catch-up vaccination for unvaccinated pregnant women and their male partners, to reduce both horizontal and vertical transmission of HBV.

Daily alcohol consumption was independently associated with increased odds of HBV infection. This association aligns with evidence from Tanzania linking alcohol use to higher HBV seroprevalence among pregnant women [36]. Alcohol may increase transmission risk through associated risky sexual behaviour and reduced uptake of preventive services. Routine alcohol screening and brief interventions should therefore be integrated into couple-based ANC and HBV prevention programmes.

In the multivariable analysis, a history of road traffic accident (RTA) showed a borderline inverse association with HBV positivity (aOR 0.20, 95% CI 0.04-1.02, p=0.05). Participants who reported RTA were less likely to test positive for HBV, although the confidence interval included values consistent with no association. We speculate that this finding may reflect greater interaction with emergency healthcare services following an RTA, potentially creating opportunities for incidental HBV screening and vaccination. However, this interpretation remains highly speculative. In Northern Uganda, where boda-boda-related injuries are common [37,38], integrating rapid HBV testing into post-trauma protocols could still be explored as a strategy to improve case detection in hard-to-reach populations, though further research is needed to confirm any relationships.

### Study limitations

This study had a few limitations. The cross-sectional design prevents causal inference between the identified factors and HBV seropositivity. Although the SD BIOLINE HBsAg rapid test has excellent reported sensitivity and specificity, it may have missed low-level or occult infections that would be detected by more sensitive ELISA or HBV DNA PCR methods and also does not detect acute or chronic infection. Some variables (alcohol consumption, vaccination history, sexual behaviour) relied on self-report and are therefore susceptible to recall and social-desirability bias. We did not assess HBeAg status, viral load, or HBV genotypes, which are important for transmission risk assessment. Despite these limitations, the study provides novel couple-level data from a high burden setting.

### Conclusions

This hospital-based study demonstrates a high HBV seroprevalence of 8.3% (40/484) among couples attending antenatal care at Gulu Regional Referral Hospital, with significantly higher rates among male partners 10.7% (n=26/242) than pregnant women 5.8% (n=14/242) with the concordance rates of 87.6%(n=212/242) and discordance rates of 12.4%. Male sex, no prior vaccination, and daily alcohol consumption were independently associated with HBsAg positivity. These findings underscore the urgent need to move beyond pregnant woman only screening. We recommend immediate implementation of routine couple HBV screening and counselling as a standard component of ANC services in Uganda, in full alignment with Ministry of Health guidelines. Male partners should be actively invited and offered onsite testing, vaccination, and linkage to care. Premarital HBV screening programmes for young people should be strengthened through community outreach and integration into existing youth-friendly services. Public health interventions targeting modifiable risk factors particularly alcohol reduction and catch-up vaccination for unvaccinated adults should be prioritised in Northern Uganda. Larger, multi-centre, community-based studies are warranted to confirm these findings and monitor the impact of couple-based interventions on HBV elimination targets.

## Data Availability

All data are in the manuscript and/or supporting information files.

## Acknowledgement

We are deeply grateful to all the couples who participated in this study. We thank the administration and staff of Gulu Regional Referral Hospital, especially the antenatal clinic team, for their support and cooperation during data collection. We acknowledge the research assistants for their dedication and professionalism. This work was conducted as part of academic requirements at Gulu University; no external funding was received.

## Data Availability

The minimal anonymised dataset and STATA analysis code are available from the corresponding author upon reasonable request.

